# Diagnosis and Tracking of SARS-CoV-2 Infection By T-Cell Receptor Sequencing

**DOI:** 10.1101/2020.11.09.20228023

**Authors:** Rachel M. Gittelman, Enrico Lavezzo, Thomas M. Snyder, H. Jabran Zahid, Rebecca Elyanow, Sudeb Dalai, Ilan Kirsch, Lance Baldo, Laura Manuto, Elisa Franchin, Claudia Del Vecchio, Monia Pacenti, Caterina Boldrin, Margherita Cattai, Francesca Saluzzo, Andrea Padoan, Mario Plebani, Fabio Simeoni, Jessica Bordini, Nicola I. Lorè, Dejan Lazarevic, Daniela M. Cirillo, Paolo Ghia, Stefano Toppo, Jonathan M. Carlson, Harlan S. Robins, Giovanni Tonon, Andrea Crisanti

## Abstract

In viral diseases T cells exert a prominent role in orchestrating the adaptive immune response and yet a comprehensive assessment of the T-cell repertoire, compared and contrasted with antibody response, after severe acute respiratory syndrome coronavirus 2 (SARS-CoV-2) infection is currently lacking. A prior population-scale study of the municipality of Vo’, Italy, conducted after the initial SARS-CoV-2 outbreak uncovered a high frequency of asymptomatic infected individuals and their role in transmission in this town. Two months later, we sampled the same population’s T-cell receptor repertoire structure in terms of both diversity (breadth) and frequency (depth) to SARS-CoV-2 antigens to identify associations with both humoral response and protection. For this purpose, we analyzed T-cell receptor and antibody signatures from over 2,200 individuals, including 76 PCR-confirmed SARS-CoV-2 cases (25 asymptomatic, 42 symptomatic, 9 hospitalized). We found that 97.4% (74/76) of PCR confirmed cases had elevated levels of T-cell receptors specific for SARS-CoV-2 antigens. The depth and breadth of the T-cell receptor repertoire were both positively associated with neutralizing antibody titers; helper CD4+ T cells directed towards viral antigens from spike protein were a primary factor in this correlation. Higher clonal depth of the T-cell response to the virus was also significantly associated with more severe disease course. A total of 40 additional suspected infections were identified based on T-cell response from the subjects without confirmatory PCR tests, mostly among those reporting symptoms or having household exposure to a PCR-confirmed infection. Taken together, these results establish that T cells are a sensitive, reliable and persistent measure of past SARS-CoV-2 infection that are differentially activated depending on disease morbidity.

## Main

Coronavirus disease 2019 (COVID-19) presents with a wide range in severity, ranging from asymptomatic infection to severe illness and death. Urgency due to the pandemic has prompted efforts to characterize the antibody response in COVID-19 patients (Iyer 2020, Isho 2020, Seow 2020, Boonyaratanakornkit 2020). However, the T cell response also plays a key role in the clearance of viral infections, governing and orchestrating both cellular and humoral immunity (Altmann 2020). Recent evidence in SARS-CoV-2 has demonstrated that the T- and B-cell responses can be discordant, and in some individuals there is a T-cell response without antibody production (Oja 2020, Sekine 2020, Gallais 2020). It remains unknown whether, and for how long, prior infection with SARS-CoV-2 provides immunity against future re-infection, nor is it known how the severity of disease might influence long-term immunity (Poland 2020). In the related SARS-CoV-1 and Middle East respiratory syndrome (MERS) coronaviruses, infections elicit an enduring T-cell response with a more fleeting antibody response suggesting the likely central role of the T-cell response in SARS-CoV-2 infection (Tang 2011, Channappanavar 2014, Zhao 2017). Therefore, direct, quantitative measures of the adaptive immune response to SARS-CoV-2 infection, particularly in longitudinal samples following recovery, may offer crucial insights into immunity and more broadly of the mechanisms underlying the immune response to this virus (Altmann 2020, Peng 2020, Weiskopf 2020).

Following the first reported COVID-19 death in the municipality of Vo’, Italy, and subsequent lockdown of the entire municipality, a large study was undertaken to screen and follow the majority of the residents in that area in an unbiased manner (Lavezzo 2020). This cohort was finely characterized in terms of demography, clinical presentation, hospitalization, comorbidities, therapies, and contact network, providing an invaluable opportunity to determine infection prevalence and transmission dynamics. Two consecutive time points, one to two weeks apart, of PCR-based diagnostic tests for SARS-CoV-2 were performed on 2,900 people, identifying 80 people who were positive for the virus. Approximately 60 days after the second PCR time point, blood samples were collected from the majority of these study participants and quantitative assessment of both SARS-CoV-2-specific T cells and IgG antibody titers were performed. As the initial burst of virus-specific effector T cells and secreted antibodies from plasmablasts are likely to have subsided two months post diagnosis, this convalescent time point is appropriate to assess longer term adaptive immune memory.

### Characterizing the T-cell response to SARS-CoV-2

T-cell receptor (TCR) sequencing was performed on blood samples from 2,291 study subjects using ImmunoSEQ®. Of these subjects with available blood, 76 had a PCR-confirmed diagnosis of COVID-19, and the rest were PCR negative at both surveys. Twenty-five of these 76 had been asymptomatic, 42 had symptoms but did not require hospitalization and 9 were hospitalized. We previously identified 4,287 public TCR sequences to SARS-CoV-2 using a case/control design that included several cohorts from the United States and Europe (Snyder 2020) and validated their diagnostic accuracy in an independent US-based cohort (Figure 1A). Notably, 85% of these sequences were also present in PCR+ Vo’ cases, and 9.4% were found at an incidence of 5% or higher. Overall, the incidence of these sequences was highly correlated across cohorts, indicating that a substantial subset of public SARS-CoV-2-specific TCRs is common across distinct populations (Figure 1B). Using these public sequences and previously defined classification framework, 74 of 76 (97.4%) of PCR+ subjects had a positive T-cell test result, with substantial variation in the number of public sequences across subjects (Figure 1C). These data indicate that detectable TCR signatures are still present two months after infection across a range of COVID-19 disease severities.

**Figure 1:**
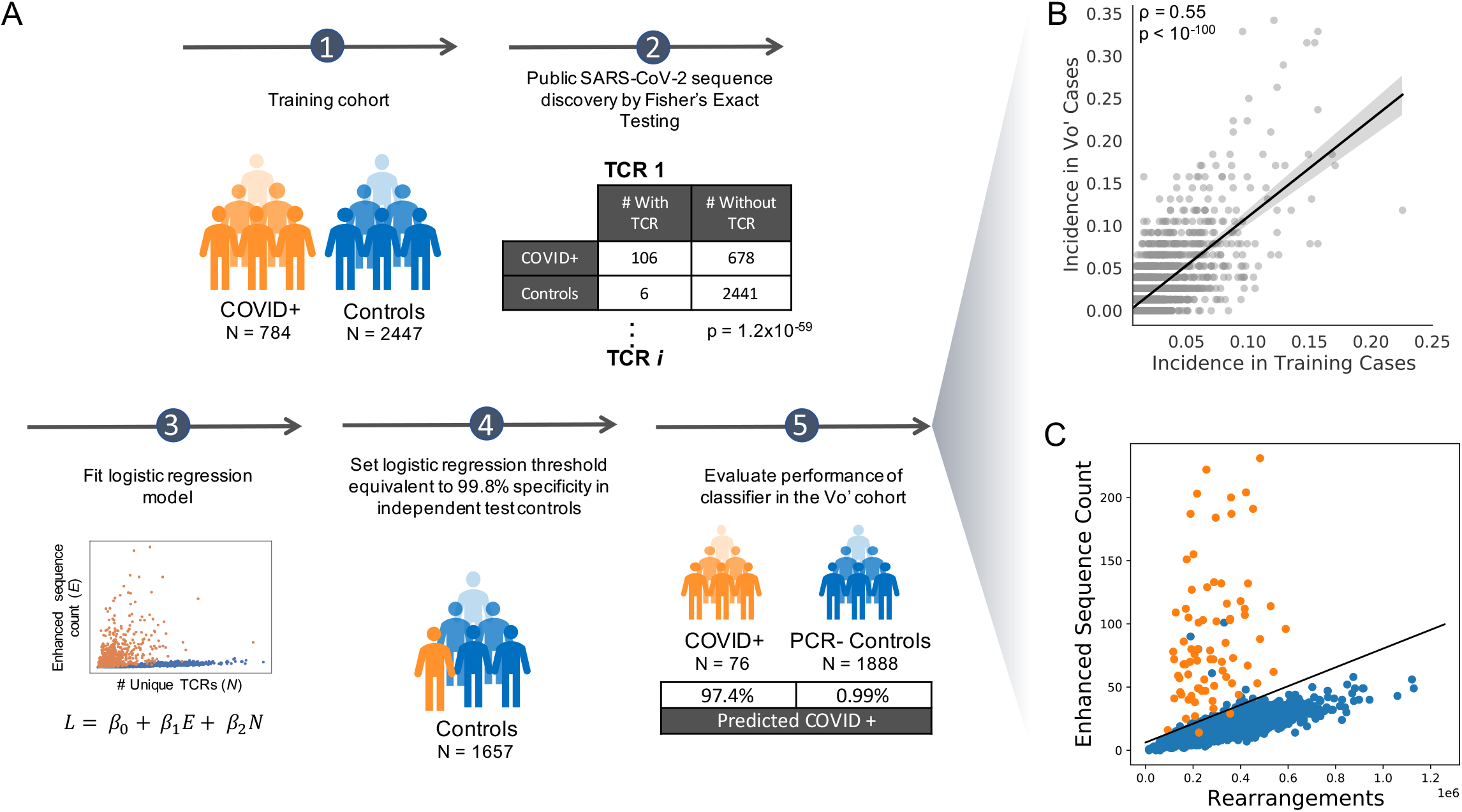
Identification and Use of Public T-Cell Receptors to Characterize the SARS-CoV-2 Immune Response. (A) Schema of general approach, starting from a case-control design and Fisher’s exact testing for each TCR on independent training data, to identify public T-cell receptor sequences that are overrepresented in cases versus controls. Following logistic regression to establish the T-cell test threshold for determining recent or past infection, the receptors are applied to this Vo’ study data set. (B) Incidence of each T-cell receptor sequence compared in the training data and in the Vo’ PCR-positive subjects. (C) The count of enhanced sequences is plotted vs. the total number of unique T-cell receptor rearrangements for subjects in the Vo’ study data set that were positive by RT-PCR (orange), or negative by RT-PCR without any additional evidence for SARS-CoV-2 infection (blue).

The two PCR+ subjects with a negative T-cell test result were both asymptomatic, indicating that the T-cell signal may vary with disease severity. To investigate this possibility, we next assessed the clonal depth and breadth of the T-cell response to SARS-CoV-2 in comparison to disease severity. Depth and breadth were calculated as defined previously (Snyder 2020), where breadth measures the relative number of distinct SARS-CoV-2-associated T cell clonotypes, and depth measures the extent to which clonotypic T cells have expanded. Clonal depth was significantly lower among individuals who reported an asymptomatic infection and increased in symptomatic and hospitalized patients (Figure 2A), while clonal breadth displayed this same trend to a lesser extent (Figure 2B). This association was present both in older (>60) and younger (≤60) subjects (Supporting Figure S1). As the clonal depth of T cells reflects the total number of cells that expanded to defend against the infection, this may be a more direct measure of response than the clonal breadth. These results are consistent with other recent findings that the magnitude of T-cell response is higher in symptomatic subjects and these differences may persist for at least 6 months (Zuo 2020), suggesting, as potential hypotheses, that differences in viral load or viral persistence during acute infection correlate both with symptoms and the depth of the T-cell response.

**Figure 2:**
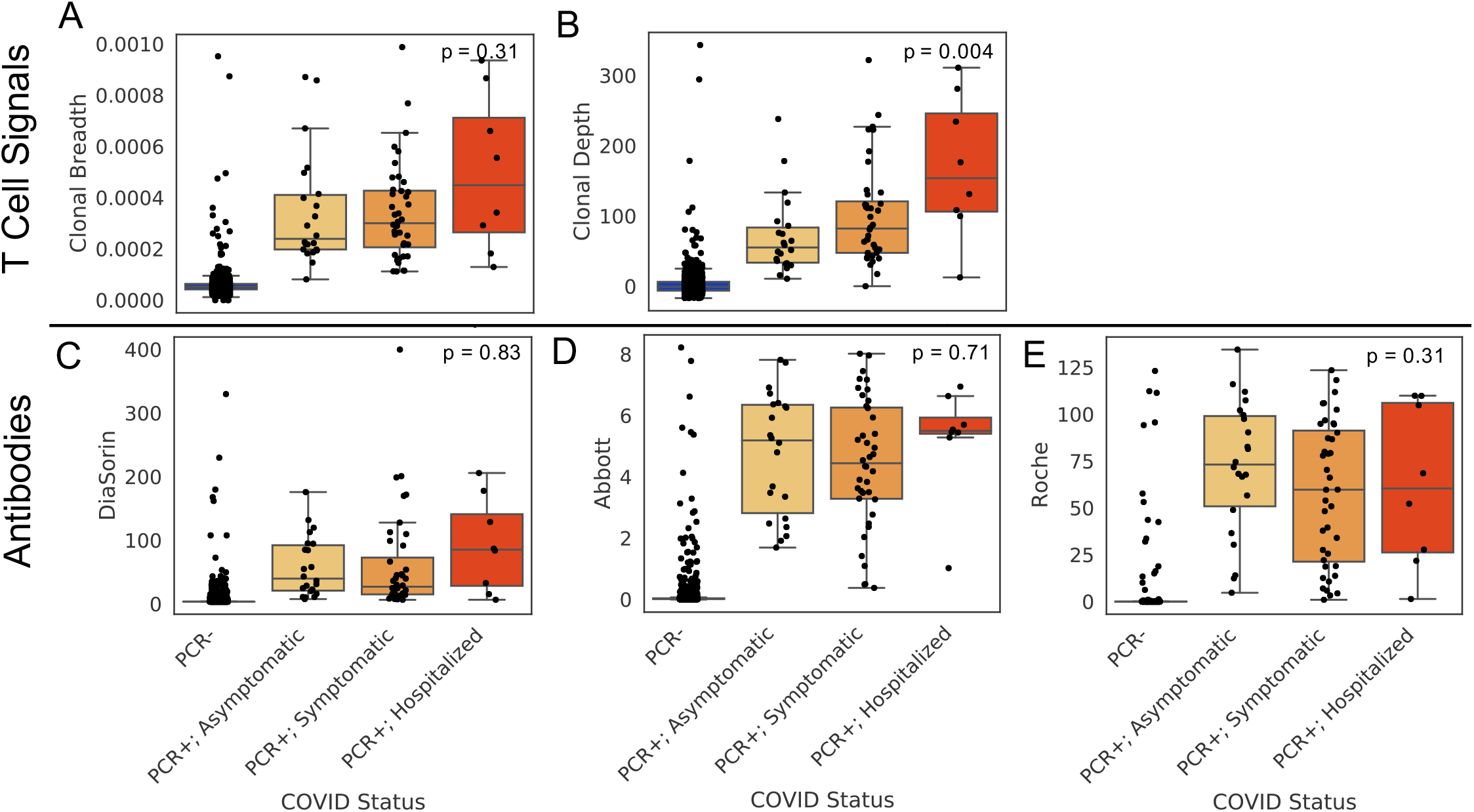
T-cell breadth (A), depth (B), and antibody levels (C-E) compared across 2154 PCR-subjects, and in 70 PCR+ subjects with results for all three serology tests faceted by disease severity. Clonal depth indicates the relative proportion of T cells that are SARS-CoV-2 specific and clonal breadth indicates the fraction of all unique TCR DNA clones that are SARS-CoV-2 specific. P values correspond to Jonckheere’s two-sided trend test across the three PCR+ categories.

### Comparisons of the T-cell and antibody responses

Antibody response (Dorigatti 2021) was measured at the same timepoint for these subjects using three commercial antibody serology tests, with 2,156 samples having results from all three assays (Supporting Table S1). Performance of these tests varied based on vendor and target. For an IgG test targeting the spike protein (DiaSorin), 78.6% (55 of 70; including 3 equivocal calls as false negatives) of PCR+ subjects were positive; the antibody negative subjects from this test were distributed relatively evenly between asymptomatic and symptomatic subjects. For two different serology tests targeting nucleocapsid phosphoprotein (IgG only from Abbott and IgG and IgM from Roche), either 65 of 70 (92.9%) or 70 of 70 (100%) of subjects were positive, similar to the T-cell test performance, which was 69 of 70 (98.6%) in this subset. In comparison to the T cell read-out, however, the observed titers of these antibodies did not correlate with severity of symptoms at 60+ days post infection for any of the assays (Figure 2C, D, E). Other studies (including Isho 2020, Iyer 2020, Long 2020, Seow 2020) have identified potential differences in antibody levels associated with severity, particularly during the acute phase of illness, but these signals decline with time and seroreversion is sometimes observed. The results here suggest that antibody signals in a longer timeframe may be less informative for measuring prior immune response, possibly due to being highly variable in the months following infection.

The extent to which T-cell measurements correspond to protective immunity remains unknown. Neutralization antibody (NAb) titers are often considered a surrogate measure of protection from infection, but are difficult to quantify reproducibly at population scale. Other antibody tests show some correlation with NAb titers (Supporting Figure S2 here, and other studies like Boonyaratanakornkit 2020), but less is known about the extent to which T-cell measures (particularly helper T cells) correlate with NAb titers. To investigate this question, neutralizing antibody titers were evaluated for all subjects who had tested positive by RT-PCR or where all four immune tests (T-cell test and three serology assays) suggested prior infection, for a total of 88 subjects. We found that both clonal depth and breadth of T cells were strongly correlated with neutralizing antibody titer (Figure 3A, B). The correlation to NAb titers was comparable to the range from serology testing (higher than the level seen for Roche and lower than that seen for DiaSorin and Abbott as in Supporting Figure S2) despite being only an indirect measure of antibody response. That two very different molecular measurements (T cells versus antibody titers) are correlated suggests that aspects of the overall adaptive immune response can be inferred using just T cells in the case of SARS-CoV-2 infection. Further research to compare T cell signals to clinical outcomes will be needed to establish their utility as a potential correlate of immunity.

**Figure 3:**
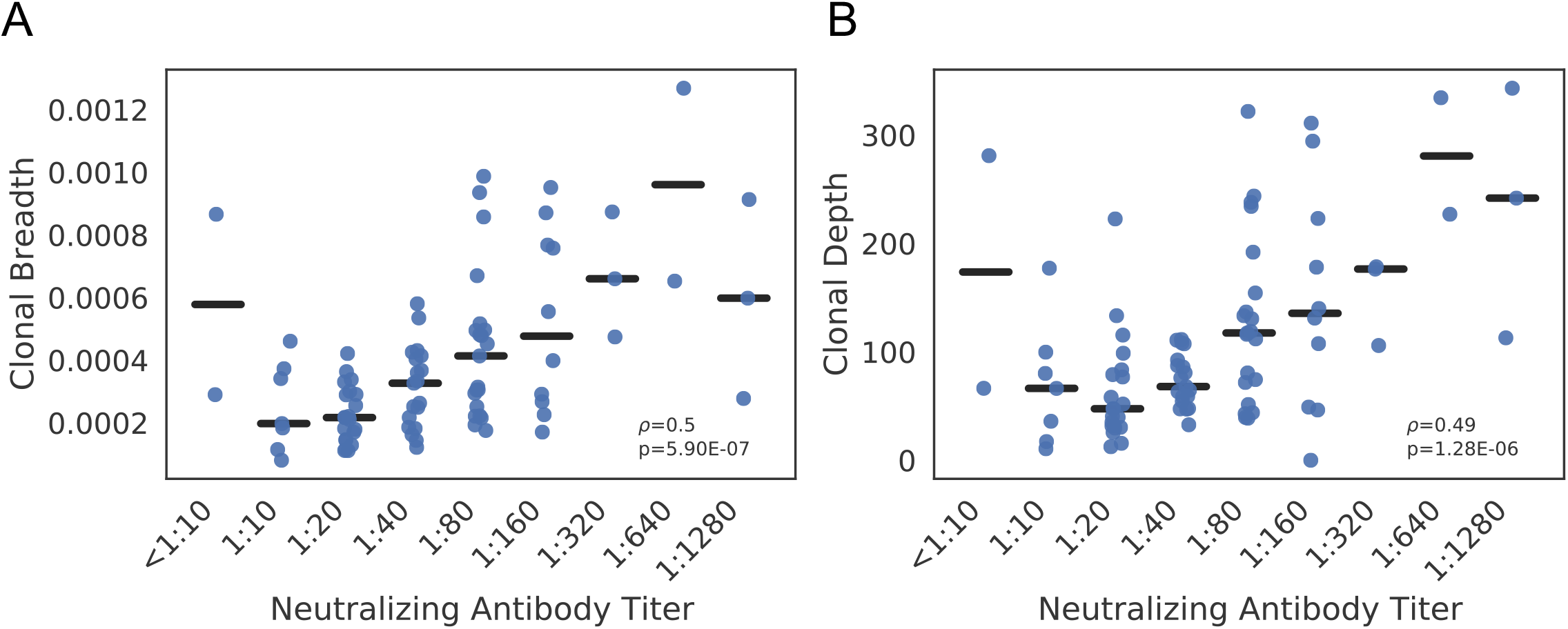
Clonal breadth (A) and depth (B) compared across 88 subjects that were positive by RT-PCR and/or all four additional tests and faceted by Neutralizing Antibody Titer. Spearman correlations are indicated by ***ρ*** and corresponding p values by p.

### CD4+ T cell responses correlate with spike-specific and neutralizing antibodies

To examine different components of the T-cell signature in more detail, we next took advantage of our prior data (Snyder 2020) to characterize the cell type and antigen specificity of the public TCR sequences used to diagnose SARS-CoV-2 infection. For many of the public TCRs and related sequences, direct observation in independent multiplexed antigen-stimulation experiments allows for assignment to a specific target (Klinger 2015). We identified 1776 and 1605 as CD8+ and CD4+ sequences, respectively, including 769 CD4+ associated sequences to spike protein and 836 CD4+ associated sequences to all other proteins.

We then compared and contrasted the breadth and depth of the CD4+ and CD8+ T cells with the B cell response. We found that both the breadth and the depth of CD4+ T cells with antigen assignments were correlated with the antibody titers, and subjects with more CD4+ T cell breadth had higher antibody levels (DiaSorin: Spearman ρ = 0.5; p = 3e-6; NAb: Spearman ρ = 0.6, p = 2e-8; Abbott: Spearman ρ = 0.5; p = 8e-6; Roche: Spearman ρ = 0.2; p = 0.03; Supporting Figure S3). Conversely, there was no correlation between CD8+ T-cell breadth and antibody levels (DiaSorin: Spearman ρ = 0.01; p = 0.92; NAb: Spearman ρ = −0.03; p = 0.8; Abbott: Spearman ρ = −0.06; p = 0.59; Roche: Spearman ρ = −0.09; p = 0.38). These results underscore the role of helper T cells in supporting the generation of antibodies, separate from the cellular immune response by cytotoxic T cells. Similar correlations were observed between antibody titers and the depth of the CD4+ T-cell response (Supporting Figure S4).

We then explored the potential association of spike-specific signal between T cells and antibodies. DiaSorin IgG spike and neutralizing antibody titers were more significantly correlated with spike-specific CD4+ T-cell breadth as compared to non-spike-specific CD4+ T-cell breadth (Supporting Figure S5). The breadth of spike-specific CD4+ T cells had a partial Spearman correlation of 0.5 (p-value = 7e-6) and 0.3 (p-value = 4e-3) to the DiaSorin IgG spike and neutralizing antibody titers, respectively. On the contrary, when we examined the association with the nucleocapsid phosphoprotein, no significant correlation between spike-specific T-cell signal and the Abbott and Roche anti-NP titers was observed. Conversely, the breadth of non-spike-specific CD4+ T cells had a partial Spearman correlation of 0.3 (p-value = 0.006) and 0.2 (p-value = 0.04) to the Abbott and Roche anti-NP titers. Overall, these results indicate that robust antibody-mediated immunity is associated with increased diversity of the antigen-specific CD4+ T cell repertoire. These results also suggest that the information obtained from the TCR repertoire data may extend beyond the direct measure of the cellular immune response and help simultaneously dissect the concomitant humoral immune response.

### Identifying additional suspected infections

The comprehensive survey across the municipality of Vo’ also allowed us to explore in detail the general population who tested negative by PCR. Notably, we identified a total of 40 (1.8%) of these 2,215, PCR-negative subjects with a positive T-cell test result, with a level of T-cell response similar to many PCR-confirmed infections (Figure 4A). Within the sampled population of PCR-negative subjects, 246 reported having some symptoms; 6.5% (16 of these subjects) had a positive T cell test. We next explored whether PCR-negative individuals residing in the same household with an individual positive for SARS-CoV-2 infection (n= 57), demonstrated a T-cell response. This analysis revealed that 21% (12 out of 57) of PCR-negative individuals with household exposure were positive for the T-cell test. Overall, positivity rates for the T-cell response were 1.0% among individuals with no reported symptoms or household exposure compared to 39% among individuals who reported both symptoms and household exposure (Figure 4B). Importantly, of the 16 subjects that were symptomatic and had detectable T-cell responses, six were symptomatic prior to the PCR surveys, indicating they may have resolved infection by the time of initial survey. 1 out of 16 reported symptoms only after the PCR surveys while the remaining 9 subjects were symptomatic during the time of survey and may have tested falsely negative by PCR. Antibody serology results were concordant with the T-cell signature for a majority of the individuals with self-reported symptoms or household exposure (Supporting Figure S7, Supporting Table S1); additional analyses focused on serology and contact tracing in this cohort are being reported separately (Dorigatti 2021).

**Figure 4:**
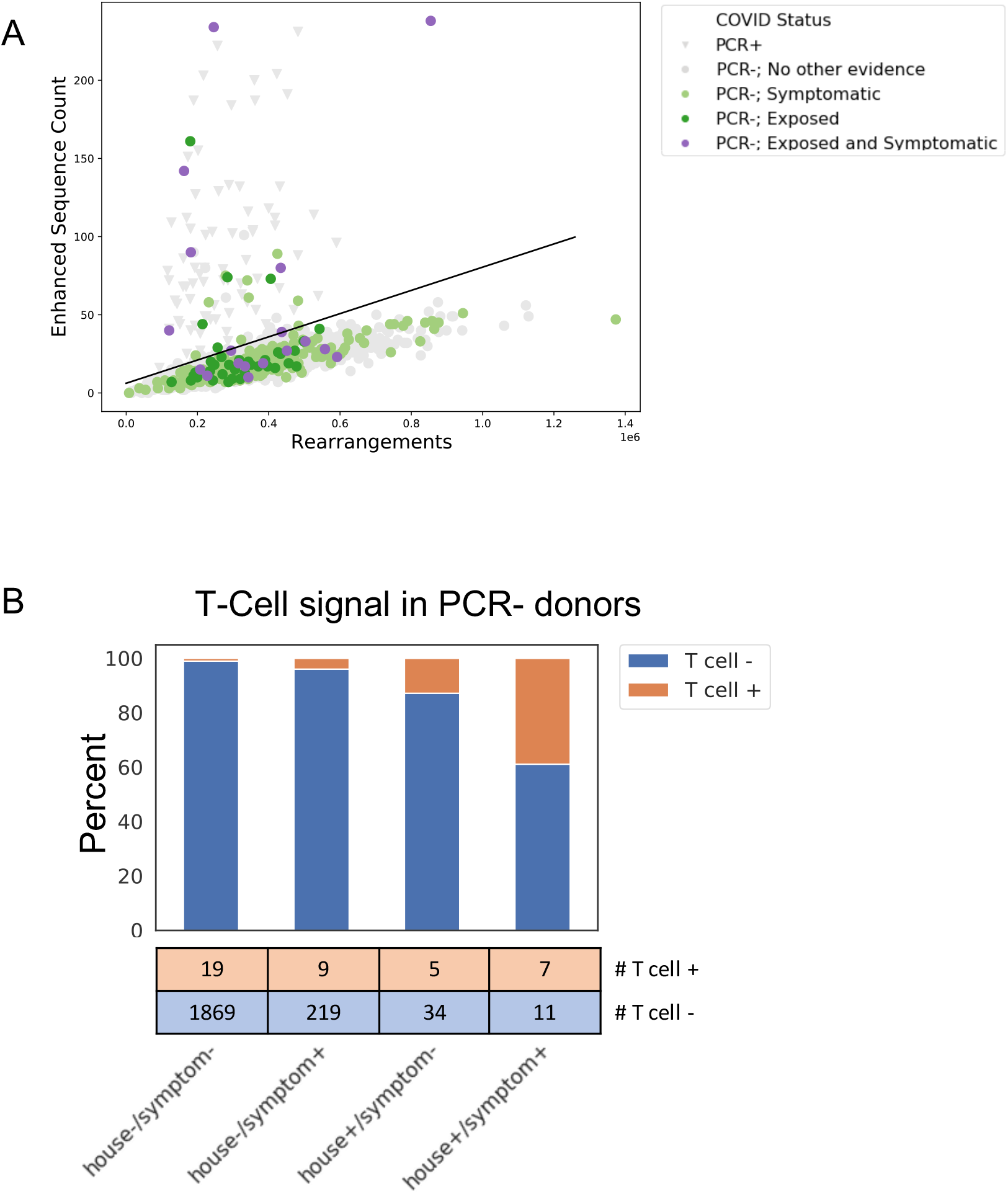
T-cell signature in RT-PCR-subjects with other evidence of SARS-CoV-2 infection. A) Count of enhanced sequences in RT-PCR-cases that were symptomatic (light green), had a household exposure (green), or were both symptomatic and had a household exposure (purple) are highlighted. RT-PCR+ (grey triangle), and RT-PCR-cases without additional evidence of SARS-CoV-2 infection (grey circle) are also shown. B) Rate of T-cell test calls in individuals with a negative RT-PCR test at the time of initial survey; positive T-cell test results were observed more often in exposed and/or symptomatic individuals.

These findings indicate that surveying the T-cell response to SARS-CoV-2 can detect cases of prior SARS-CoV-2 infection missed by PCR sampling at high sensitivity and specificity months after potential infection.

## Discussion

In this study we present evidence, gathered in an unbiased population, that the T-cell response is a highly-sensitive and specific indicator of prior SARS-CoV-2 infection. We found that T-cell responses were present at least two months after infection, even in subjects who were asymptomatic, potentially confirming prior infection in individuals who may have been exposed but did not have a confirmatory PCR test.

The findings that T-cell immunity has similar sensitivity to the antibody response in detecting SARS-CoV-2 infection have important conceptual and diagnostic implications, and yet are not entirely unexpected. Previous experiences with the related Middle East Respiratory Syndrome (MERS) and SARS-CoV-1 infections demonstrated that coronavirus-specific T cells have long term persistence and contribute to protection even in individuals without seroconversion (Tang 2011, Channappanavar 2014, Zhao 2017). Recent evidence suggests that a similar pattern is present during SARS-CoV-2 infection (Gallais 2020, Thieme 2020).

The structure of the T cell repertoire in terms of diversity and clonal expansion also provides important information to understand some of the differences that characterize symptomatic and asymptomatic individuals. Our findings demonstrate that, unlike serology tests, the T-cell response correlated with prior disease symptoms and severity at a convalescent time point over two months after infection. In addition, information gathered from the TCR repertoire analysis allow simultaneous assessment of both the cellular and humoral responses to the virus providing insight on the overall immune system response and possibly protection. The T-cell response from helper T cells strongly correlated with neutralizing antibody titer, a potential measure of protection. Helper T cells recognizing spike protein antigens correlated more strongly with the overall antibody levels to the same protein but not others, suggesting that antigen-specific CD4 T-cell help may be required for robust development of antibodies during viral infections (Sette 2008). This finding has important implications for measuring response to vaccines as well as to natural infection, since the broad set of T-cell receptors measured in this assay allows for separating responses to spike, which is the primary target of most current vaccines, from other viral proteins. The correlations between helper T cells and neutralizing antibodies may also provide new evidence relevant to disease pathology, as it has not been established whether the T-cell response is exclusively beneficial, or whether it might also contribute to immunopathology (Altmann 2020). Cases of re-infection have been described (Mulder 2020, Tillett 2020) but as they accumulate and are studied in detail, potential differences in the rates, susceptibility, or severity of re-infection may be explained by the nature of the adaptive immune response, including disease-specific memory T cells, as a measurable and reliable correlate of protection. Additional studies are imperative to further extend understanding of the role of T cells in SARS-CoV-2 infection and immunity.

## Data Availability

Clinical data for this cohort (as described in Lavezzo 2020) is available at https://github.com/ncov-ic/SEIR_Covid_Vo. T-cell repertoire profiles are available as part of the ImmuneCODE data resource (Nolan 2020), and can be downloaded from the Adaptive Biotechnologies immuneACCESS site under the immuneACCESS Terms of Use at https://clients.adaptivebiotech.com/pub/covid-2020.

https://clients.adaptivebiotech.com/pub/covid-2020

## Methods

### Clinical cohort and sample collection

This report extends results for the Vo’, Italy cohort initially described in Lavezzo 2020. Upon the detection of SARS-CoV-2 in a deceased resident of Vo’ on 21 February 2020, an epidemiological study was conducted to investigate the prevalence of SARS-CoV-2 infection in the municipality. Sampling for viral PCR testing was performed on the majority of the population immediately after the detection of the first cases (21-29 February 2020) and again at the end of a 2-week lockdown (7 March 2020). Follow-up serum and whole blood samples were collected 56 days later in early May for antibody serology and T-cell testing. Antibody response was measured using three commercial serology testing kits: LIAISON SARS-CoV-2 S1/S2 IgG (DiaSorin), Elecsys Anti-SARS-CoV-2 assay (Roche Diagnostics) and ARCHITECT SARS-CoV-2 IgG (Abbott). Serum samples were also used to perform micro-neutralisation assays; more detail on the antibody testing methods is in Dorigatti 2021.

In addition to biospecimen collection, clinical data was collected for each study participant including the results of SARS-CoV-2 testing, demographics, health records, and residence and contact network information. The definition of symptomatic used in this study is a participant who required hospitalization and/or reported fever (yes/no or a temperature above 37 °C) and/or cough and/or at least two of the following symptoms: sore throat, headache, diarrhoea, vomit, asthenia, muscle pain, joint pain, loss of taste or smell, or shortness of breath. Symptomatic subjects who reported hospitalization are split out separately as “hospitalized” in the disease severity analyses.

### Immunosequencing of T-cell receptor repertoires

Genomic DNA was extracted from frozen, plasma-depleted blood samples using the Qiagen DNeasy Blood Extraction Kit (Qiagen). As much as 18 μg of input DNA was then used to perform immunosequencing of the CDR3 regions of TCRβ chains using the ImmunoSEQ Assay. Briefly, input DNA was amplified in a bias-controlled multiplex PCR, followed by high-throughput sequencing. Sequences were collapsed and filtered to identify and quantitate the absolute abundance of each unique TCRβ CDR3 region for further analysis as previously described (Robins 2009, Robins 2012, Carlson 2013). In order to quantify the proportion of T cells out of total nucleated cells input for sequencing, or T cell fraction, a panel of reference genes present in all nucleated cells was amplified simultaneously (Pruessmann 2020).

### Characterization of the T-cell response

Classification of prior infection with SARS-CoV-2 as well as the clonal depth and breadth of T-cell response were calculated using a method similar to prior work (Snyder 2020). Briefly, T-cell receptor repertoires from 784 unique cases of RT-PCR confirmed SARS-CoV-2 infection and 2,447 healthy controls collected before 2020 were compared by one-tailed Fisher’s exact tests to identify 4,469 public TCRβ sequences (“enhanced sequences”) significantly enriched in SARS-CoV-2 positive samples. (For clarity, all training data to identify the enhanced sequences for SARS-CoV-2 infection came from multiple other study cohorts and not the population being analyzed here.) The enhanced sequences were used to develop a classifier predicting current or past infection with SARS-CoV-2 using a simple two feature logistic regression with independent variables E and N, where E is the number of unique TCRβ DNA sequences that encode an enhanced sequence and N is the total number of unique TCRβ DNA sequences in that subject. Application of this initial clinical classifier to this study demonstrated the high sensitivity (97%) reported above.

We have since developed a method to improve specificity near the decision boundary of the logistic regression by filtering enhanced sequences that may be potential false positives. Specifically, T-cell receptors that are likely associated with CMV or with multiple antigens in different HLA backgrounds and thus not truly diagnostic of SARS-CoV-2 infection are identified by Fisher’s Exact testing on TCRβ repertoires of ∼2,000 healthy controls with available HLA genotyping and CMV serotyping data. From this list of ∼1.8M sequences, the 182 sequences that were also identified as SARS-CoV-2 enhanced sequences were removed, leaving 4,287 enhanced sequences. The two-feature logistic regression classifier was refitted to the original training data using this pruned enhanced sequence list, and a decision boundary representing 99.8% specificity on 1,657 controls was used to define the test-positive threshold used in the present study. The pruned list of enhanced sequences was also used to calculate the clonal depth and breadth using the same formulae as in (Snyder 2020).

### Antigen-specific assignment of T-cell receptors

We assign public TCRs to antigens by cross-referencing enhanced sequences identified via our case/control design with TCRs observed in multiplexed antigen-stimulation experiments, both described in prior work (Snyder 2020). To maximize the number of TCR antigen assignments, we identified a set of public TCRs from an augmented sample of repertoire data. We combine the training and validation repertoires with an additional 1143 COVID positive samples accrued since this model was developed, and include samples from this study that were identified as COVID-19 negative by our T-cell test as controls. Our final sample of repertoires consists of 1,927 cases and 4,135 controls. We identified ∼500,000 public TCRs with a FET p-value < 0.05. At this level of significance, we expect a significant fraction of the public TCRs to be false positives; however, we cross-reference this list of TCRs with a set of ∼400,000 TCRs that is independently derived from our antigen-stimulation experiments yielding 3,381 overlapping TCRs. The fraction of false positive TCRs in this overlapping set is significantly smaller. These 3,381 overlapping TCRs have protein and CD4+/CD8+ assignments determined from our antigen-stimulation experiments.

We calculate the Spearman rank correlations between antibody titers and CD4+ and CD8+ T-cell response using the Pingouin package in Python (Vallat 2018) and report the two-sided significance. We note that the spike-specific CD4+ T-cell signal is correlated with the non-spike-specific signal (Spearman ρ = 0.5, p = 2e-6; Supporting Figure S4). To disentangle the confounding correlations, we calculate partial Spearman rank correlations between spike and non-spike specific T-cell response and antibody titers and report the two-sided significance. We denote the partial correlation coefficients and p values with tildes when reporting them in the figures. We examine the CD4+ T-cell response specific to spike and all other assayed proteins. When calculating the partial correlation between the antibody titers of one test and spike specific T-cell response, we take the non-spike protein specific responses as a covariate and vice versa. The partial correlations we calculate characterize the correlation between two variables that cannot be explained by the covariates and thus is conservative.

## Data Availability

Clinical data for this cohort (as described in Lavezzo 2020) is available at https://github.com/ncov-ic/SEIR_Covid_Vo. T-cell repertoire profiles and antigen annotation data from the multiplexed antigen-stimulation experiments are available as part of the ImmuneCODE data resource (Nolan 2020), and can be downloaded from the Adaptive Biotechnologies immuneACCESS site under the immuneACCESS Terms of Use at https://clients.adaptivebiotech.com/pub/covid-2020.

## Acknowledgements

We thank the population of Vo’ for volunteering en masse to participate in this study. We would like to thank Damon May for helpful discussions, and Beryl Crossley and Mitch Pesesky for assistance reviewing the data.

## Author Contributions

A.C. conceived of the initial study and, with G.T., P.G., S.T., E.L., D.M.C., I.K., T.M.S., L.B., J.M.C., H.S.R., helped design this follow-up investigation. E.L., L.M., and S.T. performed the clinical and serology data collection and curation. I.K. and H.S.R. oversaw generation of the T-cell receptor sequencing data. E.F., C.D.V., M.P., C.B., M.C., F.Sa., A.P., M.P., F. Si., J.B., N.I.L., D.L. performed laboratory testing. R.M.G., T.M.S., H.J.Z., R.E. performed data analysis. S.D., L.B. contributed to the discussion and interpretation of the clinical data. T.M.S., J.M.C., H.S.R., and G.T. wrote the manuscript, with contributions from R.M.G., S.D., I.K., E.L., S.T., D.M.C., P.G., A.C. All authors commented on and approved the final version of the manuscript.

## Author Declarations

R.M.G., T.M.S., R.E., S.D., I.K., L.B. have employment and equity ownership with Adaptive Biotechnologies. H.S.R. has employment, equity ownership, patents, and royalties with Adaptive Biotechnologies. H.J.Z. and J.M.C. have employment and equity ownership with Microsoft.

## Additional Information

Supplementary Information is available for this paper. Correspondence and requests for materials should be addressed to Dr. Crisanti, Dr. Tonon and Dr. Robins.

## Extended Figure/Table Legends

**Supporting Figure S1:**
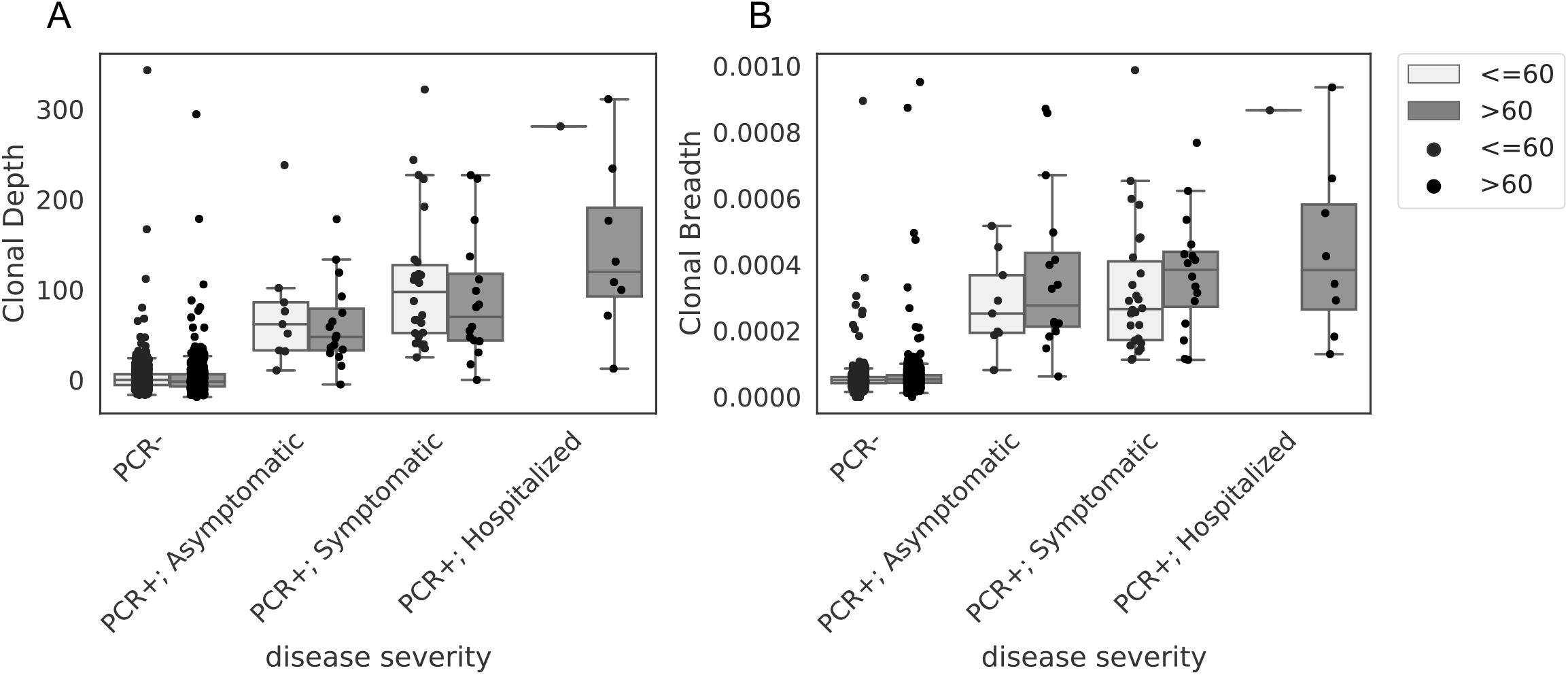
T-cell signatures stratified by disease severity and age. PCR-samples without additional evidence of SARS-CoV-2 infection are plotted alongside PCR+ samples stratified by disease severity, and age categorized as ≤60 and >60.

**Supporting Figure S2:**
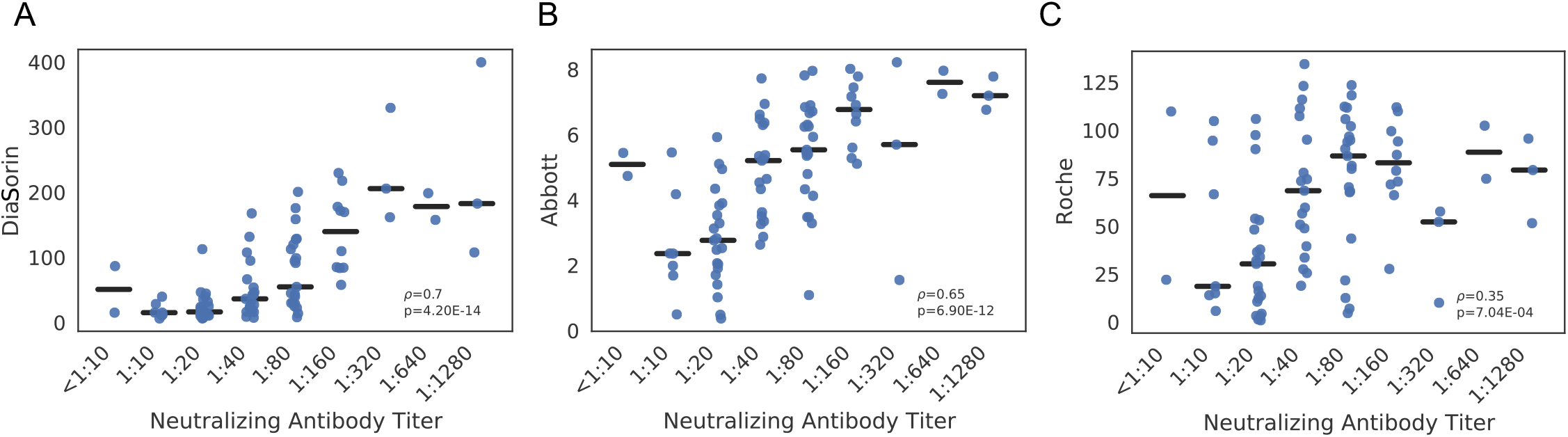
Quantitative serology scores for DiaSorin (A), Abbott (B) and Roche (C) are compared across 88 subjects and faceted by Neutralizing Antibody Titer. Spearman correlations are indicated by ***ρ*** and corresponding p values by p.

**Supporting Figure S3:**
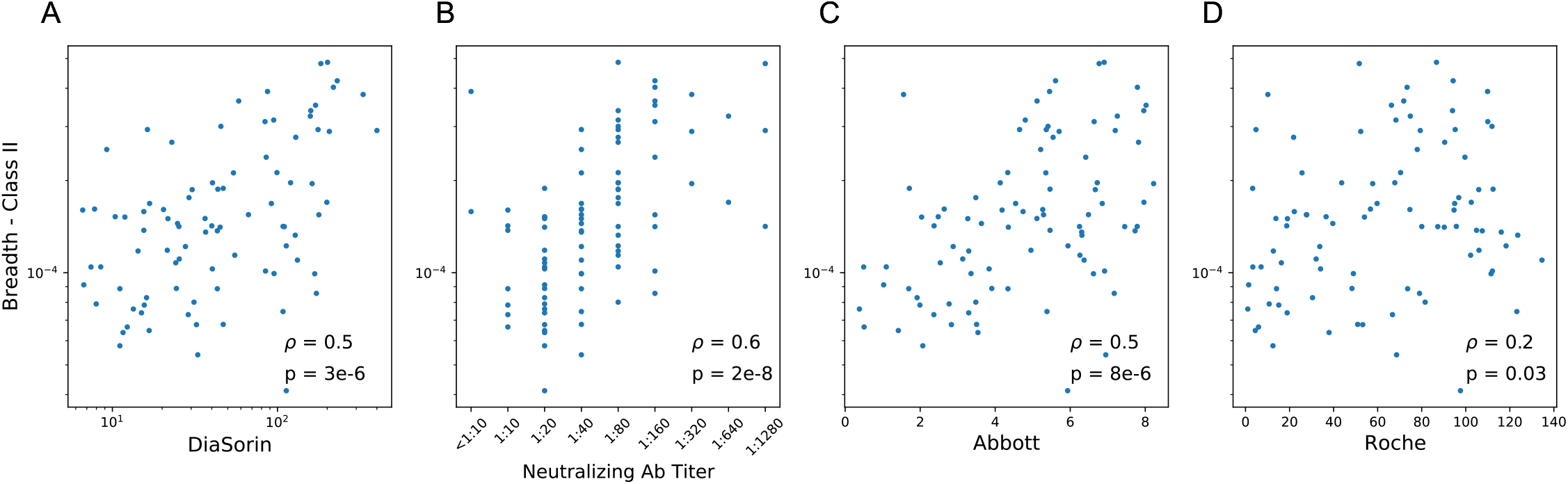
Correlation between CD4+ T cell breadth and four antibody tests. Spearman correlations are indicated by ***ρ*** and corresponding p values by p.

**Supporting Figure S4:**
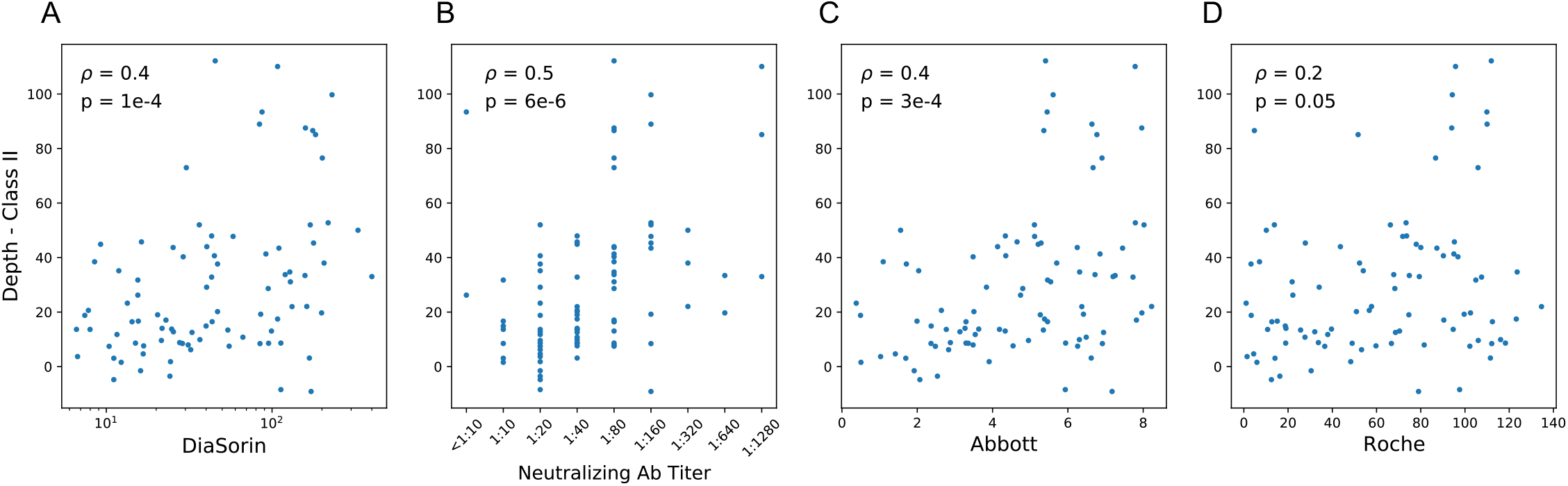
Correlation between CD4+ T cell depth and four antibody tests. Spearman correlations are indicated by ***ρ*** and corresponding p values by p.

**Supporting Figure S5:**
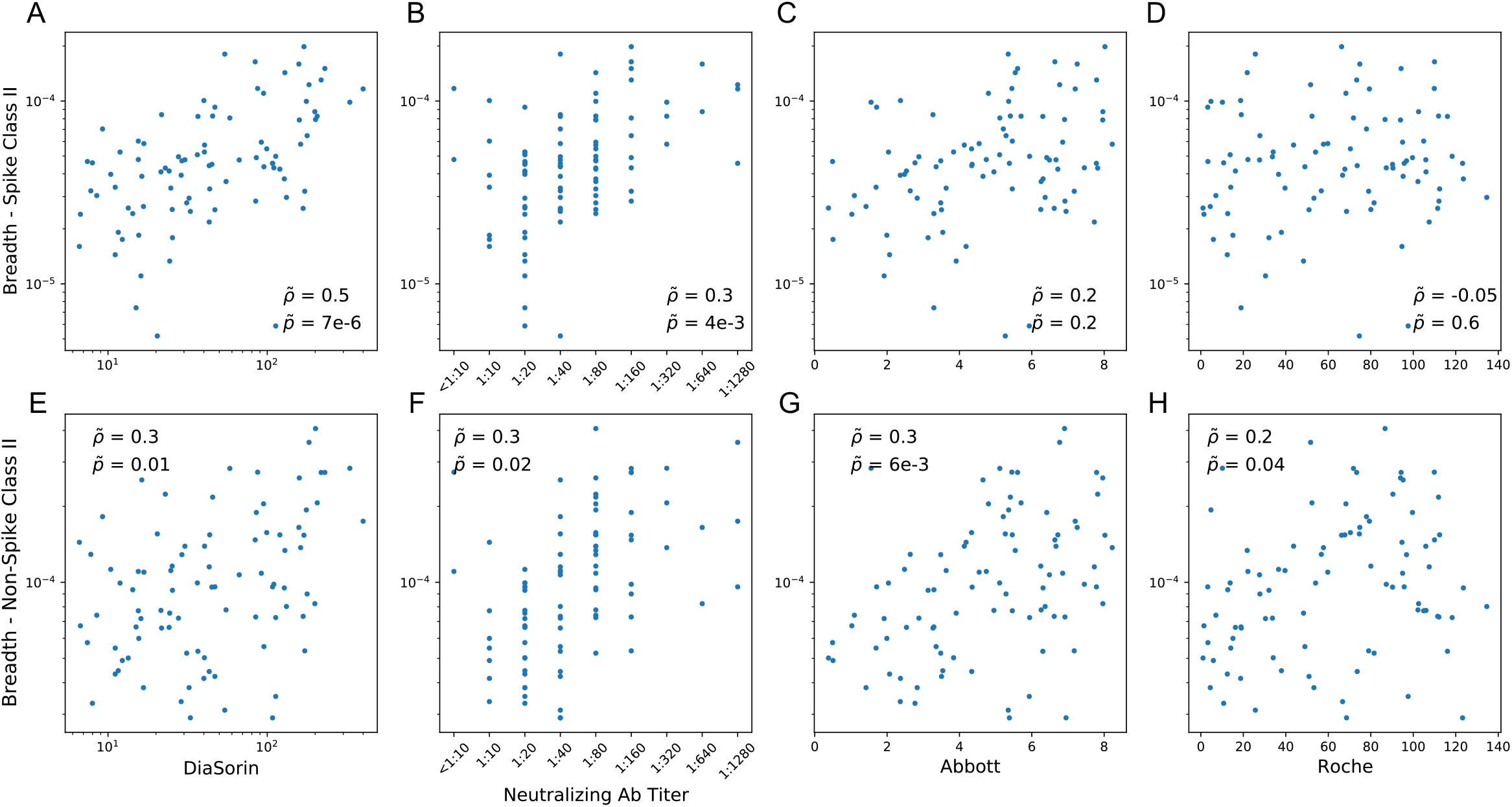
Correlation between spike-specific and non-spike CD4+ T cell breadth and antibody signals. Partial correlation analysis was performed across two T-cell sequence sets (Class II spike sequences and all other identified antigen-specific sequences) as well as four antibody tests (DiaSorin and neutralizing antibody titer for spike protein, and Abbott and Roche for nucleocapsid phosphoprotein).

**Supporting Figure S6:**
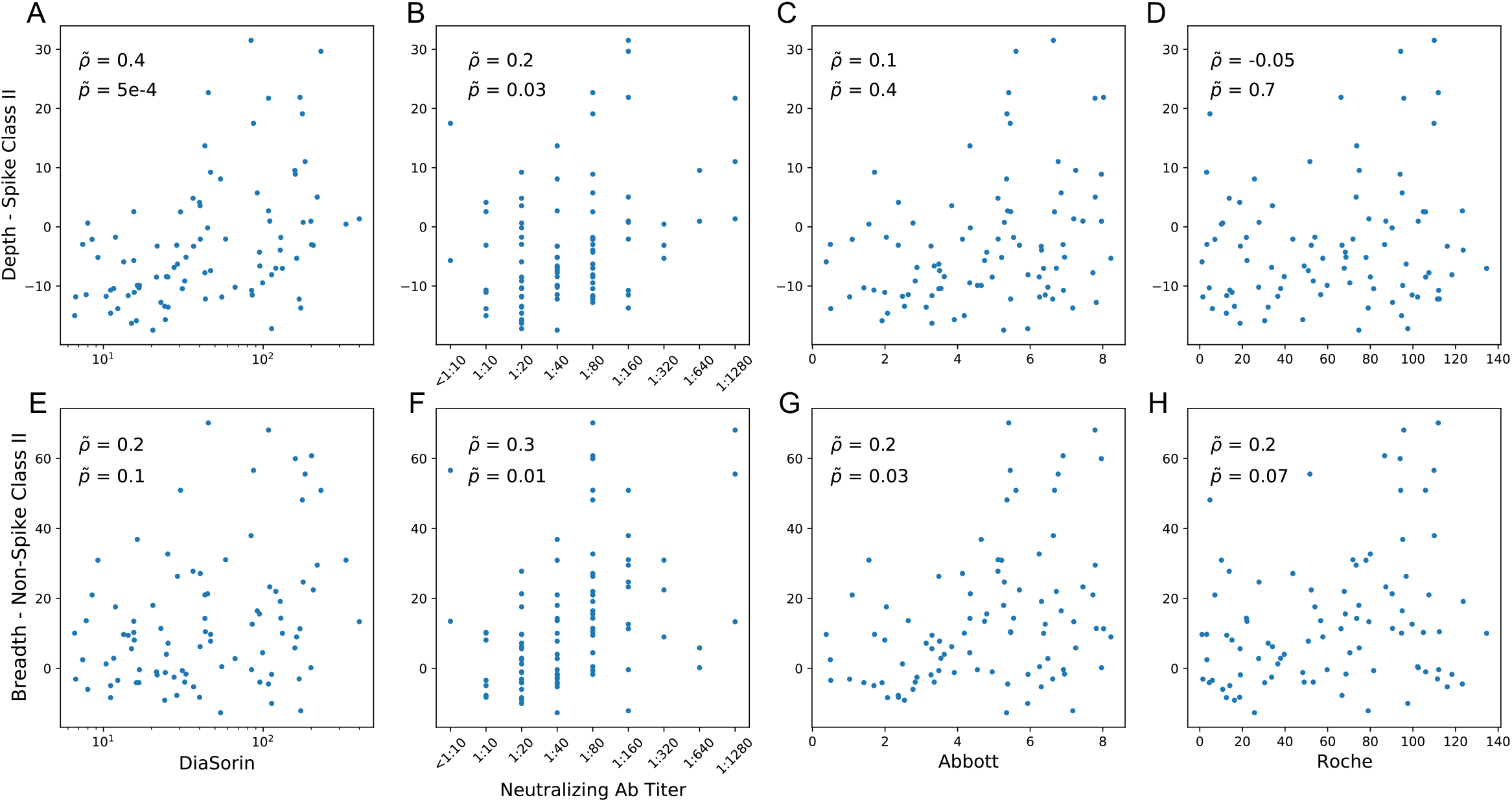
Correlation between spike-specific and non-spike CD4+ T cell depth and antibody signals. Partial correlation analysis was performed across two T-cell sequence sets (Class II spike sequences and all other identified antigen-specific sequences) as well as four antibody tests (DiaSorin and neutralizing antibody titer for spike protein, and Abbott and Roche for nucleocapsid phosphoprotein).

**Supporting Figure S7:**
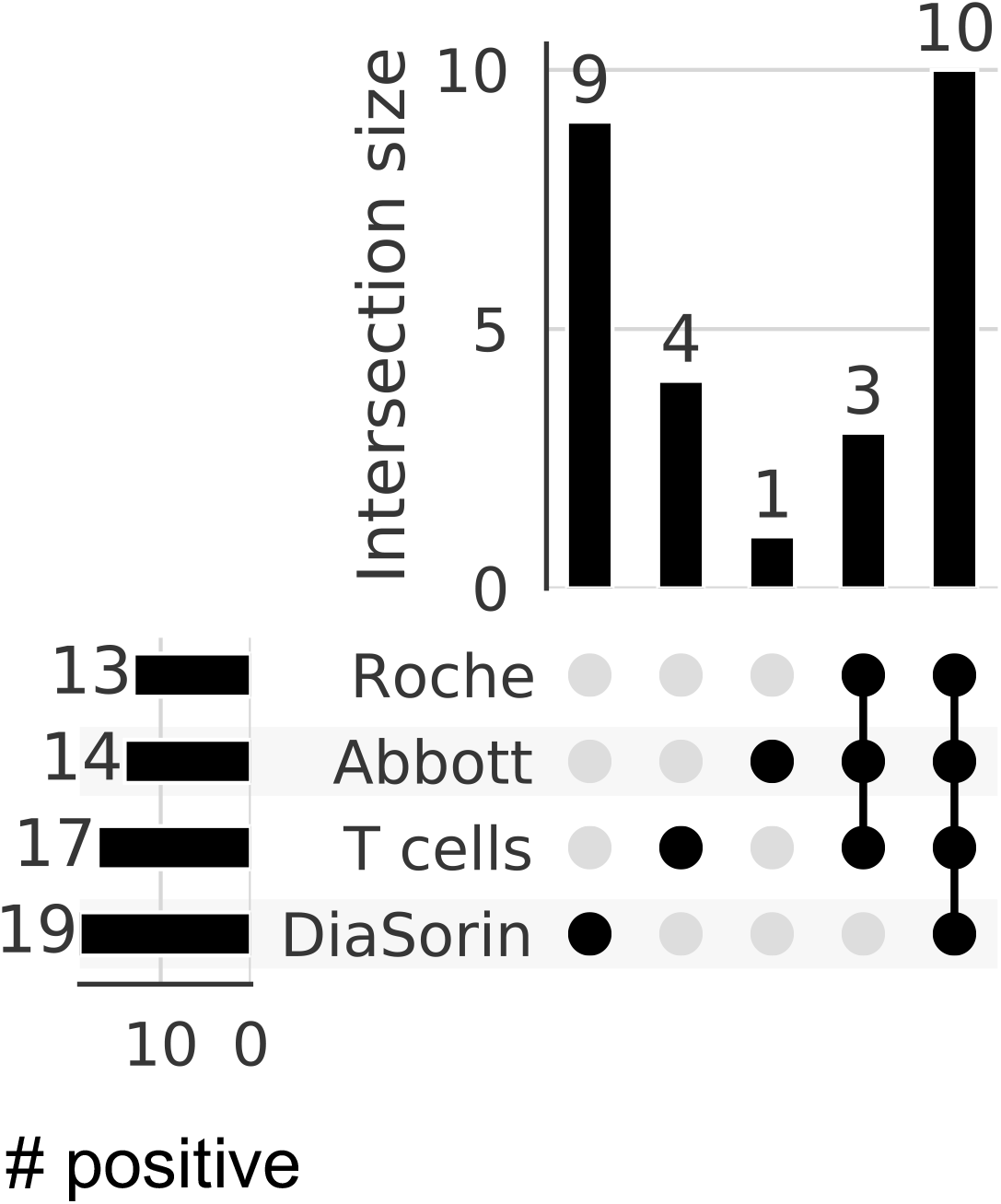
Positive T-cell and antibody tests results in PCR negative subjects with exposure and/or symptoms. In this upset plot, circles indicate overlap of each subset of among tests. For instance, 10 samples were positive by all four tests.

**Supporting Table S1:**
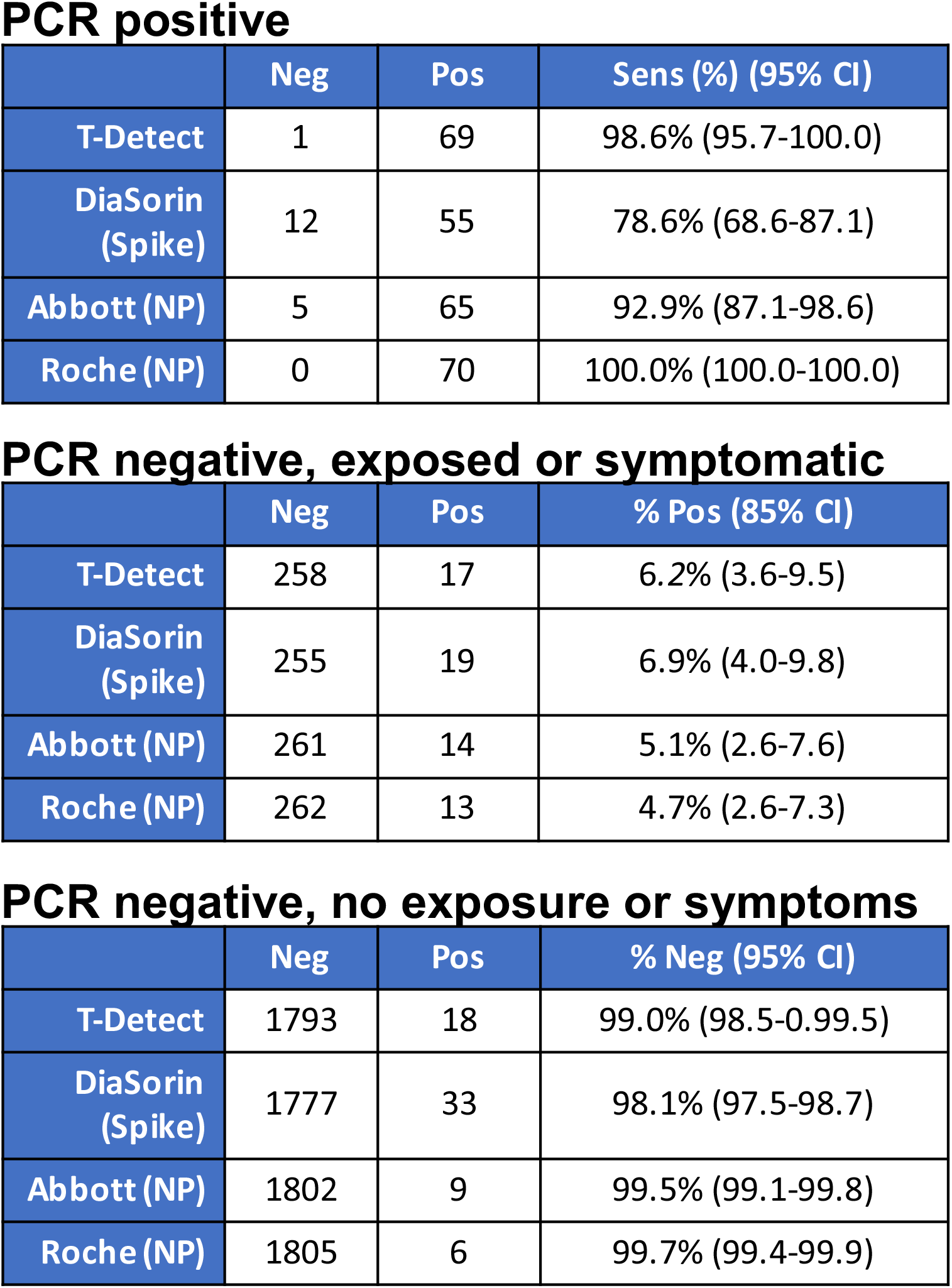
Comparison of T-cell and serology test results in three subject groups: confirmed PCR+, PCR-who share a household with a PCR+ individual and/or reported symptoms, and PCR-without household exposure or symptoms. Data represent only the samples with data from all antibody assays and T-Detect; three DiaSorin calls in the PCR positive group were equivocal and included in sensitivity estimate as false negatives. 95% confidence intervals are calculated via bootstrapping.

PCR positive
|  | Neg | Pos | Sens (%) (95% CI) |
| --- | --- | --- | --- |
| T-Detect | 1 | 69 | 98.6% (95.7-100.0) |
| DiaSorin (Spike) | 12 | 55 | 78.6% (68.6-87.1) |
| Abbott (NP) | 5 | 65 | 92.9% (87.1-98.6) |
| Roche (NP) | 0 | 70 | 100.0% (100.0-100.0) |

PCR negative, exposed or symptomatic
|  | Neg | Pos | % Pos (85% CI) |
| --- | --- | --- | --- |
| T-Detect | 258 | 17 | 6.2% (3.6-9.5) |
| DiaSorin (Spike) | 255 | 19 | 6.9% (4.0-9.8) |
| Abbott (NP) | 261 | 14 | 5.1% (2.6-7.6) |
| Roche (NP) | 262 | 13 | 4.7% (2.6-7.3) |

**Supporting Table S1: Comparison of T-cell and serology test results in three subject groups:** confirmed PCR+, PCR- who share a household with a PCR+ individual and/or reported symptoms, and PCR- without household exposure or symptoms.
|  | Neg | Pos | % Neg (95% CI) |
| --- | --- | --- | --- |
| T-Detect | 1793 | 18 | 99.0% (98.5-0.99.5) |
| DiaSorin (Spike) | 1777 | 33 | 98.1% (97.5-98.7) |
| Abbott (NP) | 1802 | 9 | 99.5% (99.1-99.8) |
| Roche (NP) | 1805 | 6 | 99.7% (99.4-99.9) |

